# Protocol for the development and evaluation of a tool for predicting risk of short-term adverse outcomes due to COVID-19 in the general UK population

**DOI:** 10.1101/2020.06.28.20141986

**Authors:** Julia Hippisley-Cox, Ash K. Clift, Carol Coupland, Ruth Keogh, Karla Diaz-Ordaz, Elizabeth Williamson, Ewen M. Harrison, Andrew Hayward, Harry Hemingway, Peter Horby, Nisha Mehta, Jonathan Benger, Kamlesh Khunti, David Speigelhalter, Aziz Sheikh, Jonathan Valabhji, Ronan A. Lyons, John Robson, Calum Semple, Frank Kee, Peter Johnson, Susan Jebb, Tony Williams, David Coggon

## Abstract

**Introduction:** Novel coronavirus 2019 (COVID-19) has propagated a global pandemic with significant health, economic and social costs. Emerging emergence has suggested that several factors may be associated with increased risk from severe outcomes or death from COVID-19. Clinical risk prediction tools have significant potential to generate individualised assessment of risk and may be useful for population stratification and other use cases.

**Methods and analysis:** We will use a prospective open cohort study of routinely collected data from 1205 general practices in England in the QResearch database. The primary outcome is COVID-19 mortality (in or out-of-hospital) defined as confirmed or suspected COVID-19 mentioned on the death certificate, or death occurring in a person with SARS-CoV-2 infection between 24^th^ January and 30^th^ April 2020. Our primary outcome in adults is COVID-19 mortality (including out of hospital and in hospital deaths). We will also examine COVID-19 hospitalisation in children. Time-to-event models will be developed in the training data to derive separate risk equations in adults (19-100 years) for males and females for evaluation of risk of each outcome within the 3-month follow-up period (24^th^ January to 30^th^ April 2020), accounting for competing risks. Predictors considered will include age, sex, ethnicity, deprivation, smoking status, alcohol intake, body mass index, pre-existing medical co-morbidities, and concurrent medication. Measures of performance (prediction errors, calibration and discrimination) will be determined in the test data for men and women separately and by ten-year age group. For children, descriptive statistics will be undertaken if there are currently too few serious events to allow development of a risk model. The final model will be externally evaluated in (a) geographically separate practices and (b) other relevant datasets as they become available.

**Ethics and dissemination:** The project has ethical approval and the results will be submitted for publication in a peer-reviewed journal.

**Strengths and limitations of the study:**

- The individual-level linkage of general practice, Public Health England testing, Hospital Episode Statistics and Office of National Statistics death register datasets enable a robust and accurate ascertainment of outcomes
- The models will be trained and evaluated in population-representative datasets of millions of individuals
- Shielding for clinically extremely vulnerable was advised and in place during the study period, therefore risk predictions influenced by the presence of some ‘shielding’ conditions may require careful consideration

## Introduction

The first cases of infection caused by the novel coronavirus SARS-CoV-2 (the virus causing coronavirus disease 2019 [COVID-19]) in the UK were confirmed on 24^th^ January 2020 and the first UK death on 28^th^ Feb 2020. Since then, the disease has spread rapidly through the population. There are currently no licensed vaccines, preventative or curative treatments for COVID-19, although one disease modifying treatment has shown promise in terms of reducing time to symptom resolution^1^. Therefore, the UK government has used social (physical) distancing and ‘shielding’ as non-pharmacological population-level interventions to limit the rate of increase in cases and adverse outcomes.

Case series of individuals with confirmed COVID-19 have identified age^2^, sex^3^, certain co-morbidities^2,4^, excess weight ^2^ and ethnicity^5^ as potentially important risk factors for exposure to infection, susceptibility to infection, hospitalisation, or death due to infection. As illustrated by a recent systematic review^6^, prediction models for COVID-19 are quickly entering the academic literature to support medical decision making at a time when they are urgently needed. Three models have been identified that predict hospital admission from pneumonia and other events (as proxy outcomes for COVID-19 pneumonia) in the general population. Ten prognostic models for predicting mortality risk, progression to severe disease, or length of hospital stay. The systematic review indicated that proposed models are poorly reported, with high risk of bias, and their reported performance is probably optimistic^6^.

Current government guidance for COVID-19 identifies individuals based on three broad levels of clinical risk (GPs are coding these as low, medium and high risk):

- Clinically Extremely Vulnerable (CEV) group: who are advised to shield (GP code: high risk)
- Clinically Vulnerable (CV) group: who are advised to follow stringent social distancing measures (GP code: medium risk)
- The remainder of the population: who are currently following mandatory social distancing measures (lockdown followed by gradual easing of measures), but who have not been advised to follow specific clinical advice (GP code: low risk).

These policies allow the ability for the National Health Service (NHS), working across government, to deliver sustained behavioural and social interventions to protect patients based on clinical need. At the heart of these policies is nationwide health data and, in this proposal, we seek to further use NHS health data to inform risk stratification. Whilst shielding and stringent social distancing are both interventions designed to reduce the risk of exposure to SARS-CoV-2, the classification of risk relates to risk of complicated or fatal disease if infected and not the risk of becoming infected.

Clinically Vulnerable (CV): Senior clinicians and expert groups in government initially identified a vulnerable group (of around 18 million), broadly identified as the at-risk group for influenza, who were eligible for a flu vaccine either on grounds of age (70 and over), pregnancy or comorbidity. 70+ was chosen rather than 65+, recognising the sharp rise in representation of the over 70s rather than the over 65s in extant hospital case series. There was an estimated 5% reduction in cases and a 15-35% reduction in mortality. On 16^th^ March 2020, the UK government recommended that this group follow stringent social distancing measures to reduce their risk of contracting the virus.

Clinically Extremely Vulnerable (CEV): Further clinical consensus emerged that, within the wider vulnerable group, a smaller number of individuals with particular conditions might be at much higher risk of complicated or fatal disease if infected. Work therefore commenced to identify a much smaller clinically extremely vulnerable group (CEV) for inclusion on a Shielded Patient List (SPL) for whom advice included staying at home and avoiding all face-to face contact. The SPL is a dynamic list, but currently sits at approximately 2.1m individuals, who are entitled to a wider government support package. This policy was announced on 22nd March 2020.

It should be noted that the advice to the Clinically Vulnerable (CV) group was not mandatory and predated wider societal measures brought in with ‘lockdown’, which now essentially sees everybody mandated to follow more stringent advice. This has resulted in the above three separate groups: CEV (who are advised to shield, also non mandatory), CV (who are advised to follow stringent social distancing measures) and the remainder of the population (who are currently following stringent social distancing measures due to lockdown, but who have not been advised to follow specific clinical advice).

Based on the best available evidence at the time, CEV patients were identified using 4 methodological approaches (described as Groups 1-4 in profession-facing communications). This approach was mirrored as far as possible by the devolved administrations:

- Method (Group) 1: Identification of a core group of CEV patients who have been identified in England by NHS Digital and contacted centrally by NHSE. This group comprised 6 categories of conditions.
- Method (Group) 2: Identification of people in particular medical subspecialties in secondary care not identifiable centrally.
- Method (Group) 3: Identification of additional CEV patients by secondary care specialists using clinical judgement.
- Method (Group) 4: Identification of a small number of CEV patients in primary care using clinical judgement.

The original SPL was intended to identify people with particular conditions which put them at highest clinical risk of severe morbidity or mortality from COVID-19, based on our understanding of the disease at the time. It was developed early in the outbreak when there were very little data or evidence about the groups most at risk of poor COVID-19 outcomes, and so was intended to be a dynamic list that would adapt as our knowledge of the disease improved and more evidence became apparent.

Given the emerging evidence on risk factors for COVID-19, and as lockdown is eased, the government is reviewing the evidence on which the initial decision to stratify patients by risk into vulnerability groups was based. This is especially relevant given that age, sex, BMI, ethnicity and certain co-morbidities are now emerging as signals for poor clinical outcomes.

As a result, England’s Chief Medical Officer asked NERVTAG to convene this expert subgroup to consider the data and evidence for risk factors for severe COVID-19 outcomes and death, and assess whether a predictive risk algorithm can be developed with the above evidence to permit a more sophisticated ‘risk stratification’ approach. There are a variety of potential uses for such a tool, but it is primarily anticipated that it could be used both clinically in informing patients of their individual risk category and managing them accordingly, and strategically to stratify the population for policy purposes.

The use of primary care datasets with contemporaneous linkage to relevant registries such as death records, hospital admissions data and COVID-19 testing results represents a novel approach to clinical risk prediction modelling for COVID-19 as they provide accurately coded, individual-level data for large numbers of a representative national population. This scenario predicates rich phenotyping of individuals with regards to a multitude of demographic, medical, pharmacological and other candidate predictors across various aspects of the health system for the purposes of robust statistical modelling, and the unparalleled sample size facilitates rigorous evaluation of any derived models. Such linked datasets have an established track record for the development, evaluation and transparent reporting of clinical risk models for conditions including (but not limited to) cardiovascular disease, diabetes and cancers ^7-11^. Therefore, the biases and limitations identified in extant risk models relevant to COVID-19 may be ameliorated by such an approach.

Any risk stratification model will need to be kept under regular review to evaluate performance and will need updating as data and disease epidemiology evolve.

### Objectives and Use Cases

In this study, we will describe the development and evaluation of novel COVID-19 risk prediction equations and corresponding tools for initial use in the NHS that will also be available internationally, subject to external evaluation per individual geography. It is anticipated that the equations will be widely available for use and that the equations will be updated regularly as understanding of COVID-19 increases, as more and better data become available, and as underlying susceptibility/immunity or policy/behaviour in the population changes or the virus itself mutates. It is also important to recognise at the outset that there will be limitations to any model that is produced and that the use of the model should be reviewed and updated regularly to ensure it remains fit for purpose.

It is also important to emphasise that this study will produce an estimated risk of both catching the virus and subsequently being seriously harmed – it will need to be further adapted to produce a risk of harm, given infection.

It is important for the population, patients, staff and the NHS that there is one widely used and appropriately developed tool which is consistently implemented across the service and which is supported by the academic, NHS and patient communities. This will then help ensure consistent policy and clear national communication between policy makers, professionals and the public.

There are numerous use cases which need to be articulated which will affect the design of the tool, but once developed, the algorithm can be adapted and developed a semi-automated fashion.

The risk algorithms can be used in various ways (examples below are based on the various ways which www.qrisk.org has been implemented and used across the NHS over the last 12 years).

- Within a consultation between the patient and a clinician (either in primary care, or the 111 service) with the intention of sharing the information with the patient to assess management options.
  - For example, a 54-year old Indian man with diabetes wishes to know his risk of serious COVID-19 in order to take action to reduce his exposure to being infected. This could be achieved through development of a risk calculator for use within a consultation.
- To electronically risk-stratify populations to appropriately target clinical or non-pharmacological interventions towards different groups of patients based on levels of risk.
  - For example, a GP practice needs to identify patients to recommend shielding, to prioritise for preventative therapies (such as weight management) or vaccination (when available). This could be achieved through the implementation of the risk equations via software embedded in GP clinical computer systems. This will ensure the tool can be applied to up-to-date electronic health records for direct clinical care purposes. It is not intended that this tool is used to make treatment decisions following admission to hospital however, since it will be developed using a general primary care population and different clinical variables will need to be considered at hospital admission. Furthermore, it does not aim to predict the expected outcomes of patients under different treatments.
- To inform mathematical modelling of the potential impact of interventions or changes in policy (e.g. shielding, prioritisation for vaccination, occupational health, health economic analyses).
  - For example, national and local governments need to assess the impact of changing guidelines on the risk categories or thresholds e.g. how many patients would be reclassified as high/medium/low risk and what the resource implications would be.
- Adapted for use by the general public to improve communication and understanding of risk through implementation into web-based tools: this would require additional segmentation in identifying, say, ‘very low risk’ populations
  - For example, a school or community needs to highlight predictors of adverse outcomes and link to recommendations in behaviours in particular to individuals at high risk to help reduce transmission of COVID-19, or individual members of the public to be better informed about the risks of different groups.
- To inform a personalised discussion between an employee and their employer regarding their occupational duties.
  - For example, key workers, health or care workers at high risk may wish to discuss working in a setting or role that has a lower risk of encountering patients with COVID-19 infection.
- Use by researchers to help generate new knowledge or insights.
  - For example, a risk stratification tool could be used to identify high risk patients to be invited to join a clinical trial or to adjust an analysis for baseline predictors of the outcome.

The initial prediction tool is being developed for public health purposes rather than for individual therapeutic or health care decisions, except for interventions aimed at prevention of infection.

## Methods

### Study design and data source

We will undertake a cohort study in a large population of primary care patients using the QResearch^®^ database (version 44). We will include all practices in England who had been using their EMIS computer system for at least a year. We will randomly allocate three quarters of QResearch practices to the training dataset and the remaining quarter to a test dataset. We will further evaluate the derived model using a dataset derived from data from GP practices using a different clinical system, ideally with a different geographical distribution from QResearch since this provides a stronger test of external validity. Further evaluations will be undertaken in external datasets to assess the models in more diverse settings. For example, we will seek to validate the algorithm using other datasets in Wales and Scotland.

### Data sources

#### QResearch-Linked database

QResearch is a high-quality research database established in 2002 which has been used extensively for the development of risk prediction tools which are widely used across the NHS^7-11^ as well as a wide range of high impact epidemiological research^12^. QResearch is a large, representative, validated GP practice research database nationally^13^. Until April 2020, there were 1205 practices contributing, covering a population of 10.5 million patients. Following a recruitment invitation, the database has now doubled to 2519 practices in England, 193 in Northern Ireland and 3 in Scotland which will cover approximately 21 million current patients. There are currently no practices in Wales.

The QResearch database is linked at an individual patient level to hospital admissions data (including intensive care unit data), COVID-19 RT-PCR positive test results held by Public Health England, the national shielded patient list held by NHS Digital, cancer registrations (including radiotherapy and systemic chemotherapy records) and mortality records obtained from the Office for National Statistics. The records are linked using a project-specific pseudonymised NHS number. The recording of NHS numbers is valid and complete for 99.8% of QResearch patients, 99.9% for ONS mortality records and 98% for hospital admissions records^14,15^.

Two new data linkages could be undertaken if available and information governance (IG) approved, linked to NHS numbers, which could then be pseudonymised using OpenPseudonymiser (www.openpseduonymiser.org) to assess:

1. Register of health care workers

2. The COVID-19 Clinical Information Network (CO-CIN) database on >40,000 patients hospitalised with COVID-19.

If these linkages were undertaken, the research objectives would be expanded to specifically examine the risks for health care workers which could directly inform policy regarding the creation of the shielded list and also quantify the risks experienced by health care workers to inform occupational health considerations.

#### English Datasets – OpenSAFELY

OpenSAFELY is a new secure analytics platform for electronic health records in the NHS, created to deliver urgent results during the global COVID-19 emergency. It is based on TPP SystmOne software from over 2000 general practices using TPP SystmOne software. It currently covers a population of 24 million patients’ full pseudonymised primary care NHS records. It is linked to records of hospital death from COVID-19, ITU data, and ONS death data including cause of death.

#### Scottish datasets – MRC funded EAVE II dataset

We will validate our risk score using the EAVE II (Early Assessment of COVID-19 epidemiology and Vaccine/anti-viral Effectiveness dataset. This project was triggered and expanded (with funding from the MRC), to pivot an NIHR-funded Pandemic Influenza Preparedness study (EAVE) to COVID-19. EAVE II has created a national electronic cohort though linking health data sets from general practice, prescribing, ICU, A&E, hospitalisations and virology testing using the unique Community Health Identification (CHI) number for residents of Scotland. EAVE II has collected electronic data from at least 5.3m individuals living in Scotland to study COVID-19.

#### Welsh datasets – SAIL

In Wales, the Secure Anonymised Information Linkage (SAIL) Databank, holds anonymised data from multiple health and non-health systems^16^. In response to the COVID-19 crisis, a complete population cohort (3.2m people) is being created, linking demographic, mortality, hospital, general practice, community prescribing, virus sequencing, COVID-19 test results and multiple other datasets to track the progress of the epidemic and evaluate the effectiveness of counter measures.

#### Study population

We will identify open cohorts of individuals registered with practices (that have been contributing to QResearch for over 12 months) on or after 24^th^ January 2020. We will exclude patients (approximately 0.1%) who do not have a valid NHS number. Patients will enter the cohort on 24^th^ January 2020 which is approximately five weeks prior to the first recorded COVID-19 death in the UK on 28^th^ February. The administrative censoring date is the date of most recent data for each outcome. The date of the outcome of interest will be observed for patients having the outcome. Patients who are not observed to have the outcome of interest in the will be censored at the administrative censoring date.

### Outcomes

#### For participants aged 19-100

Our primary outcome is COVID-19 mortality (including out of hospital and in hospital deaths) defined as confirmed or suspected COVID-19 as mentioned on the death certificate (ICD-10 codes U071 & UO72), or death occurring in a patient with a SARS-COV-2 confirmed infection in the period from 24^th^ January (date of first confirmed COVID-19 case in UK) to the last date up to which death data are available. Our secondary outcome is hospital admission in a patient with a SARS-COV-2 confirmed infection

#### For participants aged 0-18

For those aged under 19 years, our primary outcome is hospital admission in a patient with a SARS-COV-2 confirmed infection. Our secondary outcome is a composite measure including either COVID-19 mortality (as defined above) OR hospital admission SARS-COV-2 confirmed infection.

#### Candidate predictor variables

We will examine the following candidate predictor variables based on risk factors already identified in national guidance as defining or being associated with membership of clinically extremely vulnerable and clinically vulnerable groups; those already used in widely used risk equations^14^ (since these can be implemented rapidly); and variables highlighted in the related literature^9,11,17-20^. Currently (June 2020), the SPL 4 stage method allows additional patients to be added by their clinician in primary or secondary care. ^9,11,17-20^ The predictor list will be amended by future updates as new knowledge on emerging risk factors becomes available.

For analyses in people aged 0 to 18 years, we will identify predictor variables applicable to children. However, initial analyses show there are currently too few events to develop a separate risk calculator children so exploratory, descriptive analyses will be undertaken. Similarly, the analyses of pregnancy will also be considered separately.

### Demographic variables

1. Age (continuous variable)
2. Townsend deprivation score. This is an area-level continuous score based on the patients’ postcode^21^. Originally developed by Townsend^21^, it includes unemployment (as a percentage of those aged 16 and over who are economically active); non-car ownership (as a percentage of all households); non-home ownership (as a percentage of all households) and household overcrowding. These variables are measured for a given area of approximately 120 households, via the 2011 census, and combined to give a “Townsend score” for that area. A greater Townsend score implies a greater level of deprivation.
3. Ethnicity (9 categories)
4. Lives in a care home (nursing home or residential care)
5. Homelessness

### Lifestyle factors

6. Smoking status (non-smoker, ex-smoker, light, moderate, heavy)
7. Body mass index (continuous variable; in children, z-scores will be used)
8. Crack Cocaine and Injecting Drug Use
9. Alcohol dependence Current shielded list (https://digital.nhs.uk/coronavirus/shielded-patient-list#risk-criteria)
10. Solid organ transplant recipients who remain on long term immune suppression therapy
11. Cancer

a. people with cancer who are undergoing active chemotherapy
b. people with cancer who are undergoing radical radiotherapy for lung cancer
c. people with cancers of the blood or bone marrow such as leukaemia, lymphoma or myeloma who are at any stage of treatment
d. people having immunotherapy or other continuing antibody treatments for cancer *
e. people having other targeted cancer treatments which can affect the immune system, such as protein kinase inhibitors or PARP inhibitors *
f. People who have had bone marrow or stem cell transplants in the last 6 months, or who are still taking immunosuppression drugs
12. People on immunosuppression therapies sufficient to significantly increase risk of infection
13. People with severe respiratory disease^22^

a. Severe asthma – prescribed 3 or more prescriptions of steroids over the previous 12 months
b. Severe chronic obstructive pulmonary disease (COPD) prescribed 3 or more prescriptions of steroids over the previous 12 months
c. Cystic fibrosis or Interstitial Lung Disease or Sarcoidosis or non-CF bronchiectasis or pulmonary hypertension
14. People with rare diseases and inborn errors of metabolism that significantly increase risk of infection, such as severe combined immunodeficiency (SCID) or homozygous sickle cell
15. People who are pregnant with significant heart disease, congenital or acquired Moderate risk of developing complications from coronavirus (COVID-19) infection (categories as per current NHS guidance)
16. Chronic respiratory disease (non-severe)

a. Asthma
b. COPD (emphysema and chronic bronchitis)
c. Extrinsic allergic alveolitis
17. Chronic kidney disease

a. CKD stage 3 or 4
b. End-stage renal failure requiring dialysis
18. Heart disease

a. Congestive cardiac failure
b. Valvular heart disease
19. Chronic liver disease

a. Chronic infective hepatitis (hepatitis B or C infection)
b. Alcohol-related liver disease
c. Primary biliary cirrhosis
d. Primary sclerosing cholangitis
e. Haemochromatosis
20. Chronic neurological conditions

a. Epilepsy
b. Parkinson’s disease
c. Motor neurone disease
d. Multiple sclerosis
e. Cerebral palsy
f. Dementia - Alzheimer’s, vascular dementia, frontotemporal dementia
g. Down syndrome
21. Diabetes mellitus

a. Type 1 diabetes
b. Type 2 diabetes
22. Conditions or treatments predisposing to infection (including steroids)

a. Rheumatoid arthritis
b. Systemic lupus erythematosus
c. Ankylosing spondylitis or other inflammatory arthropathy such as psoriatic arthritis
d. Connective tissue disease (Ehlers-Danlos, scleroderma, Sjögren’s syndrome)
e. Polymyositis
f. Dermatomyositis
g. Vasculitis – giant cell arteritis, polyarteritis nodosa, polymyalgia rheumatica, Behçet’s syndrome, microscopic polyangitis Other medical conditions that we hypothesise may confer elevated risk:
23. Cardiovascular disease

a. Atrial fibrillation
b. Cardiovascular events (Heart attack, angina, stroke or TIA)
c. Peripheral vascular disease
d. Treated hypertension (hypertension and current antihypertensive treatment)
24. Hyperthyroidism
25. Chronic pancreatitis
26. Cirrhosis if not included (see above)

a. Including non-alcoholic steatohepatitis/fatty liver disease
27. Malabsorption

a. Coeliac disease
b. Steatorrhea
c. Blind loop syndrome
28. Peptic ulcer (gastric or duodenal ulcer, simple or complicated ulcer)
29. Learning disability
30. Osteoporosis (clinical diagnosis of osteoporosis or on DEXA scan)
31. Fragility fracture (hip, spine, shoulder or wrist fracture)
32. Severe mental illness

a. Bipolar affective disorder
b. Psychosis
c. Schizophrenia or schizoaffective disorder
33. Depression
34. HIV
35. Hyposplenism
36. Sickle cell disease
37. Tay-Sachs and other sphingolipidoses
38. History of venous thromboembolism
39. Tuberculosis

### Concurrent medication

40. Drugs affecting the immune response including systemic chemotherapy based on hospital data
41. Drugs affecting the immune response prescribed in primary care focusing on BNF chapter 8.2.
42. Long acting beta agonist (LABA)
43. Long acting muscarinic agonist (LAMA)
44. Inhaled corticosteroids (ICS)

All predictor variables will be based on the latest coded information recorded in the GP record prior to entry to the cohort.

### Development of the models

We will use the following steps:

1. Development of prognostic models for each outcome within the training data
2. Evaluation of predictive performance in the test data
3. Fitting the final model in the complete cohort and presentation of the final model

Separate models will be developed and evaluated for males and females aged 19-100 and aged <19.

### Development of the models using the training data

For all analyses, the time origin is 24^th^ January 2020 and the risk period of interest is from 24^th^ January up to the last date at which death data (for primary outcome in adults) or hospital admissions data are available. For the composite outcome of hospitalisation or death the risk period for the analysis is up to the minimum date at which death and hospital admission data are available. We will develop and validate evaluate the risk prediction equations using established methods^8,23-26^

The outcomes of interest are subject to competing risks. For the primary outcome of COVID-19 death, the competing risk is death due to other cases. For the secondary outcome of hospitalisation, the competing risk is death from any cause prior to hospitalisation. We will deal with competing risks in two ways.

First, we will use cause-specific Cox proportional hazards models for each outcome. Second, we will fit a single sub-distribution hazard (Fine and Gray) model for each outcome. Therefore, three cause-specific hazard models are fitted and three sub-distribution hazard models. All models are fitted separately in men and women. For the cause-specific hazard models, individuals are censored at the time at which a competing event occurs, for the sub-distribution hazard model individuals who did not have the outcome of interest are censored at the study end date including those who had a competing event. We will use second degree fractional polynomials (i.e. with up to two powers)^**31**^ to model non-linear relationships for continuous variables (age, body mass index and Townsend score). Separate fractional polynomial terms will be modelled for participants aged 0-19 years (when included, i.e. for the risk of hospitalisation models). Models will include interactions between BMI and ethnicity and interactions between predictor variables and age focussing on predictor variables which apply across the age range (asthma, epilepsy, diabetes, severe mental illness) where numbers allow.

### Handling of missing data

For all predictor variables, we will use the most recently available value at the time origin (24th January 2020). For indicators of co-morbidities and medication use, the absence of information being recorded is assumed to mean absence of the factor in question. There may be missing data in four variables due to never being recorded: ethnicity, Townsend score, body mass index and smoking status. We will use multiple imputation with chained equations to replace missing values for these variables ^**27-30**^. Prior to the imputation, a complete-case analysis will be fitted using a model containing only the continuous covariates within the training data to derive the fractional polynomial order and corresponding powers. For computational efficiency, we will use a combined imputation model for the three outcomes. ^31^ The imputation model will be fitted in the training data and will include all predictor variables along with age interaction terms, the interaction between BMI and ethnicity, the Nelson–Aalen estimators of the baseline cumulative hazard, and the outcome indicators (namely, death from COVID, other deaths, and hospitalisation). Separate imputation models will be fitted for men and women separately. We will carry out 5 imputations as this has a relatively high efficiency^29^ and is a pragmatic approach accounting for the size of the datasets and capacity of the available servers and software.

Each analysis model will be fitted in each imputed data set. In the outcome models BMI is modelled using fractional polynomial terms and there are interactions between BMI and ethnicity, and BMI and age. A passive imputation approach will be used, with the imputed BMI value being transformed using the fractional polynomial function derived in the complete case analysis and being used in the interaction term. We will use Rubin’s rules to combine the model parameter estimates and the baseline cause-specific survival estimates or baseline cumulative incidence across the imputed datasets^**32**^.

### Variable selection

We will fit models that include all predictor variables initially. If some predictor variables result in very sparse cells (i.e. with not enough participants or events to obtain point estimates and standard errors), we will combine some of these if clinically similar in nature. In addition, if variable selection is deemed necessary (e.g. is there is evidence of overfitting, such as very large estimated coefficients, or if the rich model cannot be fitted [does not converge] or the model performance is very poor), we will use penalised logistic models (specifically, LASSO) on the binary mortality outcome to screen (select) the variables (with the optimal penalty chosen via 5-fold cross-validation [CV]). It recommended that for best practice, well-established predictors and those with clinical credibility will be included and retained in the model. Currently, COVID-19 is a new disease and knowledge regarding which variables are established predictors is still accruing, therefore the clinical team members agreed a list of variables that should be included based on emerging evidence in the literature and public health recommendations that have been made. These clinically relevant predictor variables will then be retained in the LASSO models. Interaction terms will be considered as new variables. The variables with non-zero coefficients and the pre-specified relevant predictors will be included in the prediction models. This will be done on the complete cases (after standardising continuous variables and creating indicator variables for each category for categorical ones), and use the selected variables in the imputation models, and then the final model. In the event that logistic lasso with 5-fold CV proves too computationally intensive, and thus infeasible, we will do the variable selection on a subsample of the total data. The subsample will be a case-cohort sample, including all cases (for the outcome in question) plus a stratified random sample of the whole training data cohort (including the cases), with stratification by sex and age group, representing 10% of the training data. The selected model is then fitted in the complete training data.

### Risk equations

We will then combine the regression coefficients for each variable from the final models with an estimate of the baseline survivor function or baseline cumulative incidence to derive risk equations (cumulative incidence equations) for each outcome, with risk pertaining to the period from 24^th^ January to the latest date at which death or hospital admission data were available^**39**^. This will be done separately using the cause-specific hazard analyses and the sub-distribution hazard analyses. For the cause-specific hazard analyses the risk for a given outcome depends on the cause-specific hazard models for all competing causes and we will use a published formula^34^ to derive the cumulative incidence function in this case. ^33 33 33 33 31 29 32 32 32 32 33^

### Evaluation of the models

In the test data, we will fit an imputation model to enable imputation of missing values for ethnicity, body mass index and smoking status. The imputation model will exclude the outcome indicator and Nelson-Aalen terms, as the aim is to use the covariate data to obtain a prediction as if the outcome had not been observed. We will carry out 5 imputations. We will apply the risk equations for males and females obtained from the training data to the test data and calculate measures of performance.

As in previous studies^34^, we will calculate R^2^ values (explained variation where higher values indicate a greater proportion of variation in survival time explained by the model ^**35**^), Brier score as a measure of predictive performance, D statistics^**36**^ (a measure of discrimination which quantifies the separation in survival between patients with different levels of predicted risk where higher values indicate better discrimination) and Harrell’s C statistics at 4 months and combine these across datasets using Rubin’s rules. Harrell’s C statistic^37^ is a measure of discrimination (separation) which quantifies the extent to which those with earlier events have higher risk scores. Higher values of Harrell’s C indicate better performance of the model for predicting the relevant outcome. A value of 1 indicates that the model has perfect discrimination. A value of 0.5 indicates that the model discrimination is no better than chance. We will use adequate modifications of Brier and C-index for competing risk models.

We will calculate 95% confidence intervals for the performance statistics to allow comparisons with alternative models for the same outcome and across different subgroups. ^38^

We will assess calibration of the risk scores by comparing the mean predicted risks evaluated at 4 months with the observed risks by tenth of predicted risk. The observed risks will be obtained using nonparametric estimates of the cumulative incidences, obtained for men and women (and separately for adults/people aged<20 years).

We will also evaluate these performance measures in 6 pre-specified age groups (20-39;40-49; 50-59; 60-69; 70-79; 80+), 9 ethnic groups; individual disease conditions (e.g. diabetes), and within each of the 10 English regions.

### The final model

The evaluation above is conducted separately using the two approaches (cause-specific hazards and sub-distribution hazard models). The approach resulting in the best predictive performance, as judged through consideration of the measures and assessments outlined above, will be taken forward as the final model. We seek a model with high discrimination, low Brier score and good calibration. The imputation model fitted in the test data (excluding the outcome) is first refitted in the full data. The final outcome model is fitted in each imputed data set using the full data and estimates combined using Rubin’s Rules.

### Updating of the model using new data

We will update the models when the data are updated to cover a longer period. The baseline survivor function is likely to change after significant policy changes, so will be updated in future models to account for this where possible^32^. Even though it may not be possible to fully account for changes in baseline survival over time the risk scores will give a rank ordering of patients that can be used for risk stratification/identification of high-risk groups.

### Exploratory analyses

In addition, we will undertake a set of exploratory analyses after the aforementioned models are fitted. Given that the data used to derive the main models were generated during a time when shielding was in place, there may be underestimation of the predictive weights of some ‘shielded’ conditions, as we were unable to observe the ‘full potential risk’ of these diseases. To examine this further, we will use a stratified Cox model approach. Here, we will stratify the population on the basis of shielding status (a binary measure), thus allowing the baseline hazard to vary in the two sub-groups of the study cohort. When obtaining predictions for the shielded population, we will use the baseline hazard for the non-shielded sub-cohort. Another exploratory analysis may include fitting the model in different time periods – such as in the time between first positive cases and 2 weeks after the date on which individuals were first advised to shield/were added to the shielded patient list. We would also fit the model during the time period where shielding occurred and compare model performance.

### Development of risk categories

Since there is no currently accepted threshold for classifying high risk of COVID-19 outcomes, we will examine the distribution of predicted risks and calculate a series of centile values. For each centile threshold, we will calculate the sensitivity, specificity, positive and negative predictive values of the risk scores over a 4-month follow-up. A range of possible categorisations will also be explored corresponding to different levels of risk, based on the risk score derived from the log-hazard ratios (coefficients) of the primary Cox regression model.

We will use all the relevant patients on the database to maximise the power and generalisability of the results. We will use Stata (version 16) and R statistical software for analyses. We will adhere to the TRIPOD statement for reporting^39^.

### Patient and public involvement

Patients and members of the public will be involved in setting the research question, the outcome measures, the design, implementation and dissemination of the study findings. A multi-ethnic panel of patient representatives will also advise on dissemination including the use of culturally appropriate lay summaries describing the research and its results.

### Implementation

A web-based program will implement the risk-stratification procedure. In a similar manner for QRISK, a range of alternative communication formats are possible and will be empirically evaluated. These could include a full risk-score, a risk-categorisation, alternative representations of relative and absolute risk for the appropriate risk-category, or translation into a ‘COVID-age’. Multiple versions of the tool will be available to allow the user to directly enter information (for example, via an “app”) as well as versions which allow pre-population of exiting data via electronic health care record systems. In all implementations it will be made clear that the risks being communicated are not ‘your’ risks: they are essentially what we have observed based on medical records in a group of initially uninfected people.

The prediction model is for the combined risk of infection and then death (or hospitalisation). Therefore, it will be important to emphasise that the model is a prediction model and is not a causal model, and as such the model parameters do not have a causal interpretation. Despite this, there may be potential risks regarding misinterpretation of the results of our work. As such, we will work towards clear and robust communication of this in any clinical or public-facing tools, including but not limited to any publications that result from this work.

NHS Digital will ensure that implementations meet medical device regulations requirements. For transparency and to ensure wide availability of the resulting tools, all code underlying the implementation will be published as open source software and freely available.

## Discussion of methodology and approach

### Ethical considerations

We see an important distinction between (a) factors which are included in a risk equation to ensure that the risk estimates are as accurate as possible and (b) how the risk equation is then used in guidelines and clinical practice to ensure ethical, effective, and equitable access to services for everyone.

The primary purpose of our study is to report on the development and validation of new risk equations rather than to produce national policy or clinical guidance although we recognise the results may be used by policy makers and clinicians. All clinical decisions regarding the beneficial and safe use of these risk equations necessarily remain the responsibility of the attending health care professional. This initial tool is being developed for public health purposes rather than for individual therapeutic or health care decisions, except for interventions aimed at prevention of infection.

There are, however, ethical issues to consider regarding how the tools may be used. We have analysed this within the “four ethical principles” framework which is widely used in medical decision making. The four principles are autonomy, beneficence, justice and non-maleficence^40^. The new risk equations, when implemented in clinical software, are designed to provide more accurate information for patients and clinicians on which to base decisions thereby promoting shared decision making and patient autonomy. They are intended to result in clinical benefit by identifying where changes in management are likely to benefit patients, thereby promoting the principle of beneficence. Justice can be achieved by ensuring that the use of the risk equations results in fair and equitable access to health services which is commensurate with the patients’ level of risk. Lastly the risk assessment must not be used in a way which causes harm either to the individual patient or to others (for example, by introducing or withdrawing treatments where this is not in the patients’ best interest) thereby supporting the non-maleficence principle. How this applies in clinical practice will naturally depend on many factors especially the patient’s wishes, the evidence based for any interventions, the clinician’s experience, national priorities and the available resources. The risk assessment equations therefore supplement clinical decision making not replace it.

As this work is conducted, we must be aware that as COVID-19 is a poorly understood disease in terms of its clinical behaviour and risks, the outputs may contain results that seem to be counter-intuitive, or go against what we understand for other diseases. Should this occur, we will rigorously explain that any new observations in this project cannot be interpreted as being causal and should not form the basis for any change in behaviour or trigger any intervention. If novel insights or counterintuitive results are obtained, these will be explored further in other studies, as this does not fall under the remit of our aim to produce a prediction tool.

### Comparison with other risk scores

A recent systematic review by Wynants et al^6^, et al. identified 27 studies reporting 31 multivariable risk prediction models pertaining to either the prediction of hospital admission for COVID pneumonia (actual or presumed, n=3), diagnostic models for detecting infection (n=18), or prognostic models in confirmed cases (n=10). Their review also covered non-peer reviewed manuscripts submitted on open access pre-print repositories.

Whilst all included studies were identified as being at high risk of bias (as per PROBAST), and very few offered meaningful indication of the details of their construction or intended use, they offer a reference point for the status and quality of risk prediction modelling in this pandemic. Interestingly, the review did not identify any models that predicted risk of death in the general population as our study intends to do as its primary outcome of interest, the prognostic models instead estimated mortality risk in those with suspected or confirmed COVID infection (n=6), risk of prolonged hospital stay (n=2), or progression to severe disease/a critical state (n=2).

Regarding our secondary endpoint of interest (risk of hospitalisation due to COVID-19), three models were identified by Wynants, et al^6^. aiming to predict risk of hospital admission at a general population level. However, none of these models were developed using a dataset that contained any COVID-19 cases, instead using admission for a suite of respiratory infections as proxy events.

In contrast to these models that have a high risk of bias, non-representative samples and imperfectly described methodology, our study cohort covers a significant proportion of the United Kingdom and is population-representative, has the advantage of accurate ascertainment of myriad predictors and outcomes, and we will adhere to consensus reporting guidelines (TRIPOD).

### Strengths

The methods to derive and validate these models are broadly the same as for a range of other widely used clinical risk prediction tools derived from the QResearch database ^7-11^. The strengths and limitations of the approach have already been discussed in detail ^8,11,23,24,41,42^. In summary, key strengths include size, wealth of data on risk factors, good ascertainment of outcomes through multiple record linkage, prospective recording of outcomes, use of an established validated database which has been used to develop many risk prediction tools, and lack of selection, recall and respondent bias and robust analysis. UK general practices have good levels of accuracy and completeness in recording clinical diagnoses and prescribed medications ^43^. We think our study has good face validity since it has been conducted in the setting where most patients in the UK are assessed, treated and followed up.

### Limitations

Limitations of our study include the lack of formal adjudication of diagnoses, potential for misclassification of outcomes depending on testing, information bias, and potential for bias due to missing data. Our database has linked mortality and hospital admissions data and is therefore likely to have picked up the great majority of COVID-19 related ICU admissions and death thereby minimising ascertainment bias. The initial evaluation will be done on a separate set of practices and individuals to those which were used to develop the score although the practices all use the same GP clinical computer system (EMIS – the computer system used by 55% of UK GPs). An independent evaluation will be a more stringent test and should be done (e.g. using data from different clinical systems or the other countries within the UK), but when such independent studies have examined other risk equations, ^41,42,44,45^ they have demonstrated similar performance compared with the validation in the QResearch database^7,8,23^. Whilst our study population is from England and is representative of the English population, we will need to evaluate its performance in other datasets which are representative of the other devolved nations. We are however validating the risk score using both the EAVE II national cohort which includes data on over 90% of the Scottish population. A particular limitation in the COVID-19 context is that much of the outcome data will have been generated whilst social distancing and other public health measures have been in place, and therefore may not be fully representative of risk in the absence of these measures.

For transparency, we will publish the model itself and source code for the equations on a publicly available website with an appropriate license (e.g. LGPL). The rationale for this is to ensure that those interested in reviewing or using the open source will then be able to find the latest available version as the score continues to be updated.

## Conclusion

We will develop a set of new equations to quantify risk of adverse COVID-19 outcomes, taking account of demographic, social and clinical variables. The equations are intended to provide a valid measure of those outcomes the general population of patients tested in separate validation cohort. The equations can be used in conjunction clinical guidelines to enable patients to be identified for focused assessments and interventions.

## Data Availability

To guarantee the confidentiality of personal and health information only the authors have had access to the data during the study in accordance with the relevant licence agreements. Access to the QResearch data is according to the information on the QResearch website (www.qresearch.org). The equations presented in this paper will be released as Open Source Software under the GNU lesser GPL v3 to ensure transparency and wide availability. The open source software allows use without charge.

## Acknowledgements

We acknowledge the contribution of EMIS (Egton Medical Information Systems) practices who contribute to the QResearch database and EMIS, and the Universities of Nottingham and Oxford for expertise in establishing, developing, and supporting the QResearch database. QResearch acknowledges funding from the NIHR funded Nottingham Biomedical Research Centre. The data on COVID-19 PCR tests were used with permission from Public Health England. The intensive care data were used with permission from ICNARC. The Hospital Episode Statistics data and Civil registration date used in this analysis are re-used by permission from the NHS Digital who retain the copyright.

## Funding

This work will be funded by a grant from the National Institute for Health Research. QResearch receives NIHR Oxford Biomedical Research Centre; the Medical Sciences Division of the University of Oxford; John Fell Oxford University Press Research Fund; the Oxford Wellcome Institutional Strategic Support Fund (204826/Z/16/Z); Cancer Research UK (CR-UK) grant number C5255/A18085, through the Cancer Research UK Oxford Centre. It also receives contributions in kind from EMIS Health (commercial supplier of NHS health computer systems). KC is supported by the British Heart Foundation (CH/16/1/32013). RHK is supported by a UKRI Future Leaders Fellowship (MR/S017968/1) KDO is supported by a Royal Society-Wellcome Trust Sir Henry Dale Fellowship 218554/Z/19/Z). KK is supported by National Institute for Health Research Applied Research Collaboration East Midlands (NIHR ARC-EM) and the NIHR Leicester Biomedical Research Centre. AS is supported by Health Data Research UK (MR/R008345/1) and the Medical Research Council (MR/R008345/1).

## Ethical approval

The protocol for QResearch has been published in e-Prints and was reviewed in accordance with the requirements for the East Midlands Derby research ethic committee (ref 18/EM/0400). An amendment was approved on 01 April 2020 to allow data linkage with the PHE test results and TPP data.

## Declaration

The lead author affirms that the manuscript resulting from this work will be an honest, accurate, and transparent account of the study being reported; that no important aspects of the study have been omitted and that any discrepancies from the study as planned have been explained.

## Competing Interests

JHC reports grants from National Institute for Health Research Biomedical Research Centre, Oxford, grants from John Fell Oxford University Press Research Fund, grants from Cancer Research UK (CR-UK) grant number C5255/A18085, through the Cancer Research UK Oxford Centre, grants from the Oxford Wellcome Institutional Strategic Support Fund (204826/Z/16/Z), during the conduct of the study. JHC is an unpaid director of QResearch, a not-for-profit organisation which is a partnership between the University of Oxford and EMIS Health who supply the QResearch database used for this work. JHC is a founder and shareholder of ClinRisk ltd and was its medical director until 31^st^ May 2019. ClinRisk Ltd produces open and closed source software to implement clinical risk algorithms (outside the proposed work) into clinical computer systems. CC reports receiving personal fees from ClinRisk Ltd., outside this work. KK reports grants from NIHR, is the National Lead for ethnicity and diversity for the National Institute for Health Applied Research Collaborations, and Director of the University of Leicester Centre for Black Minority Ethnic Health. AH reports being a member of the New and Emerging Respiratory Virus Threats Advisory Group. PJ reports employment by NHS England during the conduct of the study, grants from Epizyme, Janssen, personal fees from Takeda, Bristol-Myers-Squibb, personal fees from Novartis, Celgene, Boehringer Ingelheim, Kite Therapeutics, Genmab, and Incyte, all outside the submitted work. AKC reports personal fees from Huma Therapeutics, outside of the scope of the submitted work. RL reports grants from Health Data Research UK outside the submitted work. AS reports grants from MRC during the conduct of the study. CS reports grants from the DHSC National Institute of Health Research UK, Medical Research Council UK, and the Health Protection Unit in Emerging and Zoonotic Infections (University of Liverpool) during the conduct of the study, and is a minority owner in Integrum Scientific LLC (Greensboro, NC, USA) outside of the submitted work. RHK, JB, KDO, JR, NM, EW, EH, HH, PH, DS, JV, FK, SJ, TW and DC have no conflicts of interests to declare.

## Notes

### Author Declarations

East Midlands - Derby Research Ethics Committee 18/EM/0400

## References

1. Ledford H. Hopes rise on coronavirus drug remdesivir. Nature 2020.

2. Garg S KL, Whitaker M, et al. Hospitalization Rates and Characteristics of Patients Hospitalized with Laboratory-Confirmed Coronavirus Disease 2019 — COVID-NET, 14 States, March 1–30, 2020. MMWR Morb Mortal Wkly Rep 2020; 69: 458–64.

3. Piva S, Filippini M, Turla F, et al. Clinical presentation and initial management critically ill patients with severe acute respiratory syndrome coronavirus 2 (SARS-CoV-2) infection in Brescia, Italy. J Crit Care 2020; 58: 29–33.

4. Wang B LR, Lu Z, Huang Y. Does comorbidity increase the risk of patients with COVID-19: evidence from meta-analysis. Aging 2020; 8;12(7):6049–6057.

5. Pareek M, Bangash MN, Pareek N, et al. Ethnicity and COVID-19: an urgent public health research priority. The Lancet 2020.

6. Wynants L, Van Calster B, Bonten MMJ, et al. Prediction models for diagnosis and prognosis of covid-19 infection: systematic review and critical appraisal. BMJ 2020; 369: m1328.

7. Hippisley-Cox J, Coupland C, Vinogradova Y, et al. Predicting cardiovascular risk in England and Wales: prospective derivation and validation of QRISK2. BMJ 2008:bmj.39609.449676.25.

8. Hippisley-Cox J, Coupland C, Robson J, Sheikh A, Brindle P. Predicting risk of type 2 diabetes in England and Wales: prospective derivation and validation of QDScore. BMJ 2009; 338: b880..

9. Hippisley-Cox J, Coupland C. Derivation and validation of updated QFracture algorithm to predict risk of osteoporotic fracture in primary care in the United Kingdom: prospective open cohort study. BMJ 2012; 344 (may22 1): e3427–e.

10. Hippisley-Cox J, Coupland C. Predicting the risk of Chronic Kidney Disease in Men and Women in England and Wales: prospective derivation and external validation of the QKidney(R) Scores. BMC Family Practice 2010; 11: 49.

11. Hippisley-Cox J, Coupland C. Development and validation of risk prediction algorithm (QThrombosis) to estimate future risk of venous thromboembolism: prospective cohort study. BMJ 2011; 343: d4656.

12. Coupland CAC, Hill T, Dening T, Morriss R, Moore M, Hippisley-Cox J. Anticholinergic Drug Exposure and the Risk of Dementia: A Nested Case-Control Study. JAMA Intern Med 2019.

13. Kontopantelis E, Stevens RJ, Helms PJ, Edwards D, Doran T, Ashcroft DM. Spatial distribution of clinical computer systems in primary care in England in 2016 and implications for primary care electronic medical record databases: a cross-sectional population study. BMJ Open 2018; 8(2).

14. Hippisley-Cox J, Coupland C. Predicting risk of emergency admission to hospital using primary care data: derivation and validation of QAdmissions score. BMJ Open 2013; 3(8): e003482.

15. Hippisley-Cox J. Validity and completeness of the NHS Number in primary and secondary care electronic data in England 1991-2013.2013. Hippisley-Cox J. Validity and completeness of the NHS number in primary and secondary care: electronic data in England 1991-2013 http://eprints.nottingham.ac.uk/3153/1/Validity%26CompletenessNHSNumber.pdf (accessed June 2013).

16 Lyons RA, Jones KH, John G, et al. The SAIL databank: linking multiple health and social care datasets. BMC Med Inform Decis Mak 2009; 9: 3.

17 Hippisley-Cox J, Coupland C. Symptoms and risk factors to identify men with suspected cancer in primary care: derivation and validation of an algorithm. Br J Gen Pract 2013; 63(606): 1–10.

18 Hippisley-Cox J, Coupland C. Symptoms and risk factors to identify women with suspected cancer in primary care: derivation and validation of an algorithm. Br J Gen Pract 2013; 63(606): 11–21.

19 Clegg A, Bates C, Young J, et al. Development and validation of an electronic frailty index using routine primary care electronic health record data. Age and Ageing 2016.

20 National Institute for Clinical Excellence. Multimorbidity: clinical assessment and management, NICE guidelines NG56. London, 2016.

21 Townsend P, Davidson N. The Black report. London: Penguin; 1982.

22 Digital N. COVID-19 identifying patients for shielding. 2020. https://digital.nhs.uk/coronavirus/shielded-patient-list/methodology/rule-logic.

23 Hippisley-Cox J, Coupland C. Predicting risk of osteoporotic fracture in men and women in England and Wales: prospective derivation and validation of QFractureScores. BMJ 2009; 339: b4229.

24 Hippisley-Cox J, Coupland C, Vinogradova Y, Robson J, Brindle P. Performance of the QRISK cardiovascular risk prediction algorithm in an independent UK sample of patients from general practice: a validation study. Heart 2008; 94: 34–9.

25 Hippisley-Cox J, Coupland C, Brindle P. Development and validation of QRISK3 risk prediction algorithms to estimate future risk of cardiovascular disease: prospective cohort study. BMJ 2017; 357: j2099.

26 Hippisley-Cox J, Coupland C. Development and validation of QDiabetes-2018 risk prediction algorithm to estimate future risk of type 2 diabetes: cohort study. BMJ 2017; 359: j5019.

27 Schafer J, Graham J. Missing data: our view of the state of the art. Psychological Methods 2002; 7: 147–77.

28 Group TAM. Academic Medicine: problems and solutions. BMJ 1989; 298: 573–9.

29 Steyerberg EW, van Veen M. Imputation is beneficial for handling missing data in predictive models. J Epidemiol Community Health 2007; 60: 979.

30 Moons KGM, Donders Rart, Stijnen T, Harrell FJ. Using the outcome for imputation of missing predictor values was preferred. J Epidemiol Community Health 2006; 59: 1092.

31 Schafer JL. Multiple imputation: a primer. Stat Methods Med Res 1999; 8(1): 3–15.

32 Rubin DB. Multiple Imputation for Non-response in Surveys. New York: John Wiley; 1987.

33 Booth S, Riley RD, Ensor J, Lambert PC, Rutherford MJ. Temporal recalibration for improving prognostic model development and risk predictions in settings where survival is improving over time. Int J Epidemiol 2020.

34 Hippisley-Cox J, Coupland C, Brindle P. The performance of seven QPrediction risk scores in an independent external sample of patients from general practice: a validation study. BMJ Open 2014; 4(8): e005809.

35 Royston P. Explained variation for survival models. Stata J 2006; 6: 1–14.

36 Royston P, Sauerbrei W. A new measure of prognostic separation in survival data. Stat Med 2004; 23: 723–48.

37 Harrell F, Lee K, Mark D. Multivariable prognostic models: issues in developing models, evaluating assumptions and adequacy, and measuring and reducing errors. Stat Med 1996; 15: 361–87.

38 Newson RB. Comparing the predictive powers of survival models using Harrell’s C or Somers’ D. Stata Journal 2010; 10(3): 339–58.

39 Collins GS, Reitsma JB, Altman DG, Moons KGM. Transparent Reporting of a multivariable prediction model for Individual Prognosis Or Diagnosis (TRIPOD): The TRIPOD StatementThe TRIPOD Statement. Annals of Internal Medicine 2015; 162(1): 55–63.

40 Gillon R. Medical ethics: four principles plus attention to scope. BMJ 1994; 309(6948): 184.

41 Collins GS, Mallett S, Altman DG. Predicting risk of osteoporotic and hip fracture in the United Kingdom: prospective independent and external validation of QFractureScores. BMJ 2011; 342: d3651.

42 Collins GS, Altman DG. External validation of the QDScore for predicting the 10-year risk of developing Type 2 diabetes. Diabetic Medicine 2011; 28: 599–607.

43 Majeed A. Sources, uses, strengths and limitations of data collected in primary care in England. Health Statistics Quarterly 2004; (21): 5–14.

44 Collins GS, Altman DG. Predicting the 10 year risk of cardiovascular disease in the United Kingdom: independent and external validation of an updated version of QRISK2. BMJ 2012; 344: e4181.

45 Collins GS, Altman DG. An independent and external validation of QRISK2 cardiovascular disease risk score: a prospective open cohort study. BMJ 2010; 340: c2442.

